# Endocrine disrupting chemicals and COVID-19 relationships: a computational systems biology approach

**DOI:** 10.1101/2020.07.10.20150714

**Authors:** Qier Wu, Xavier Coumoul, Philippe Grandjean, Robert Barouki, Karine Audouze

## Abstract

**Background:** Patients at high risk of severe forms of COVID-19 frequently suffer from chronic diseases, but other risk factors may also play a role. Environmental stressors, such as endocrine disrupting chemicals (EDCs), can contribute to certain chronic diseases and might aggravate the course of COVID-19.

**Objectives:** To explore putative links between EDCs and COVID-19 severity, an integrative systems biology approach was constructed and applied.

**Methods:** As a first step, relevant data sets were compiled from major data sources. Biological associations of major EDCs to proteins were extracted from the CompTox database. Associations between proteins and diseases known as important COVID-19 comorbidities were obtained from the GeneCards and DisGeNET databases. Based on these data, we developed a tripartite network (EDCs-proteins-diseases) and used it to identify proteins overlapping between the EDCs and the diseases. Signaling pathways for common proteins were then investigated by over-representation analysis.

**Results:** We found several statistically significant pathways that may be dysregulated by EDCs and that may also be involved in COVID-19 severity. The Th17 and the AGE/RAGE signaling pathways were particularly promising.

**Conclusions:** Pathways were identified as possible targets of EDCs and as contributors to COVID-19 severity, thereby highlighting possible links between exposure to environmental chemicals and disease development. This study also documents the application of computational systems biology methods as a relevant approach to increase the understanding of molecular mechanisms linking EDCs and human diseases, thereby contributing to toxicology prediction.

## Introduction

The COVID-19 pandemic started in the fall of 2019 and spread to a large part of the world during the winter and spring of 2020. By late June 2020, it had led to more than 500,000 deaths, of which one-fourth in the US and about 175,000 in the EU (https://coronavirus.jhu.edu/map.html, https://covid19.who.int/). Despite considerable research activities, there are still many unknowns concerning this infectious disease, especially with regard to the substantial variability of the disease severity. Following an initial infectious phase, a “cytokine storm”, leading to pneumonia is observed in severe cases which may require intensive care. It is still unclear why infections lead to severe cases in some patients and not in others, but both endogenous and exogenous factors can likely influence the outcome of the disease.

In addition to older age and male sex, several comorbidities are associated with severe COVID-19 and increased mortality risk. Disorders such as cardiovascular disease, type II diabetes (T2D), obesity, chronic respiratory disease or hypertension are strongly linked to severe COVID-19 cases (Petrilli et al. 2020)(Zhou et al. 2020)(Stefan et al. 2020). As has recently been proposed, underlying metabolic and endocrine dysfunctions may be mechanistically linked to the exacerbation of the coronavirus infection (Bornstein et al. 2020), and these observations may inspire new insight into the pathogenesis of this disease, including biological interpretation of the mechanisms involved. Environmental stressors have already been suggested to contribute to the severity of the disease (Bashir et al. 2020; Fattorini and Regoli 2020; Zhu et al. 2020), but little mechanistic support for this association is available. A relevant approach would be to compare the biological pathways triggered by environmental stressors with those involved in the COVID-19 severity. If similar pathways are found, this would increase the likelihood that such stressors may contribute to critical stages of this disease.

Given the suspected hormonal mode of vulnerability (Drucker 2020) endocrine disrupting chemicals (EDCs) could represent important triggers of aggravated infection, e.g., in the form of phthalates, bisphenols, organochlorine pesticides, and perfluorinated alkane substances (PFASs) (Trasande et al. 2016; Vandenberg et al. 2016). Exposure to these substances may affect the immune defense, thus potentially increasing the susceptibility to develop COVID-19 (Tsatsakis et al. 2020), as supported by experimental studies(Cipelli et al. 2014; Couleau et al. 2015). For example, epidemiological evidence on children exposed to PFASs show decreased immune responses to routine vaccines (Grandjean et al. 2012) and a greater risk of developing infectious disease(Dalsager et al. 2016; Granum et al. 2013).

As promising tools to gain better insight into the possible risk factors and mechanisms, toxicological and chemical data sources have expanded substantially, thereby enabling network science and computational systems biology methods to become feasible (Audouze et al. 2013, 2018; Taboureau and Audouze 2017; Vermeulen et al. 2020; Wu et al. 2020). We have therefore conducted an integrative systems biology exploration to identify overlapping proteins that are both dysregulated by EDCs and involved in comorbidities associated with aggravated COVID-19. Based on this tripartite network, integrating protein-EDC associations and protein-disease annotations, we then performed biological enrichments of pathways to detect the most plausible relationships between EDC exposure and COVID-19 severity.

## Methods

We employed a computational systems biology approach to explore putative linkages between EDCs and COVID-19 as presented in Figure 1. First, a tripartite network was created based on known associations between proteins and either COVID-19 comorbidities or EDCs, as compiled from existing databases (CompTox, DisGeNET, GeneCards) (A). Then, biological enrichment was performed with the jointly identified proteins (i.e., those retrieved in both association studies) (B) by over-representation analysis (ORA) to identify the pathways that were the highly linked to both the diseases and the EDCs (C). As a final step, the biological pathways were explored with available knowledge regarding COVID-19 mechanisms (from the literature and the AOP-Wiki database), thereby allowing consideration of hypothetical linkages between EDCs and COVID-19 (D).

**Figure 1.**
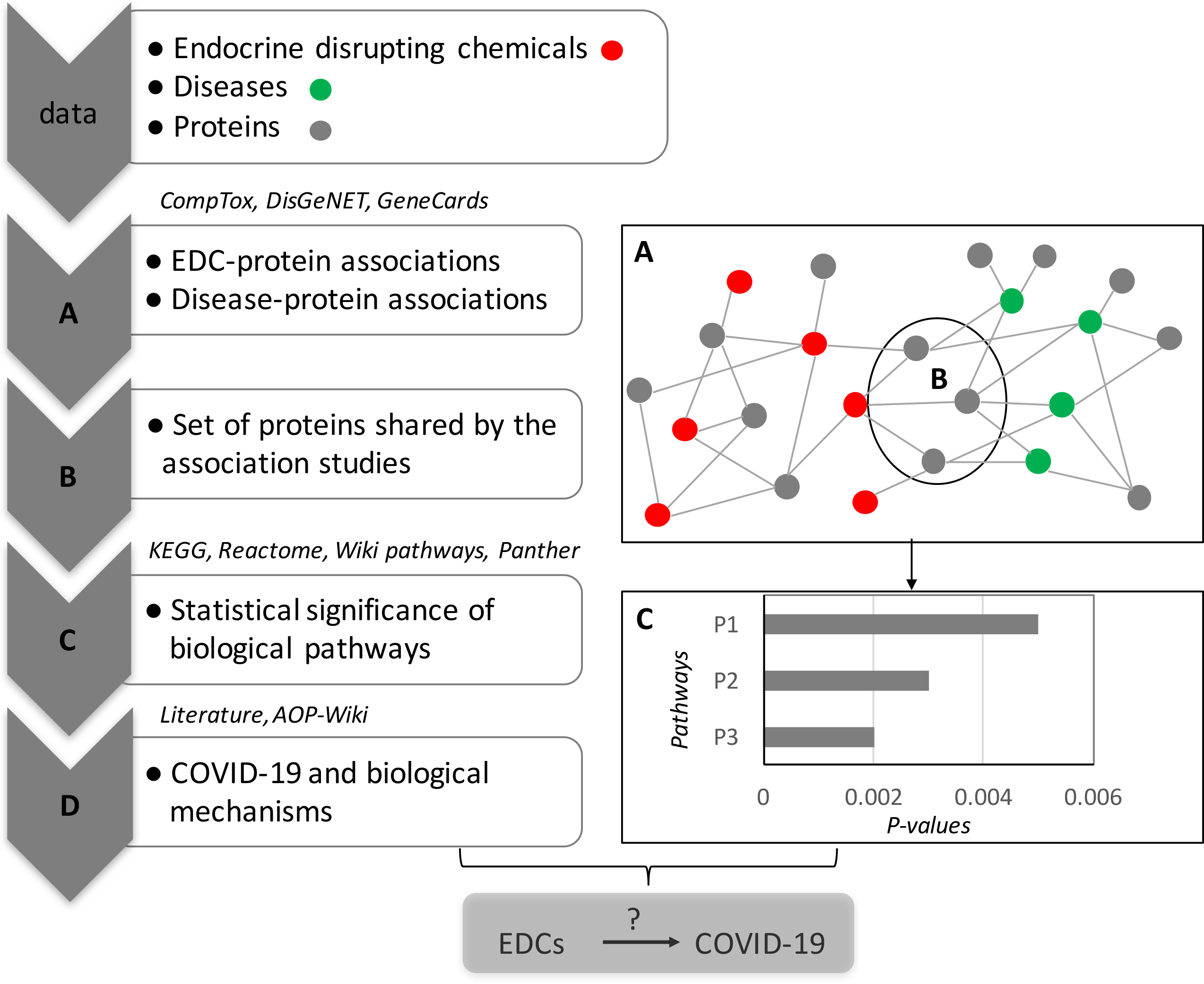
Overview of the integrative systems toxicology approach. **A**: Human proteins known to be dysregulated by endocrine-disrupting chemicals (EDCs) were extracted from the CompTox database; human proteins linked to obesity or to comorbidities or metabolic dysfunction known to be associated with obesity were compiled using DisGeNET and GeneCards. These compiled data were used to develop a tripartite network. **B:** A set of proteins was identified that was common to both association studies (proteins targeted by the EDCs and also involved in comorbidities). **C**: Biological enrichment was performed for pathways for each of the four databases, by over-representation analysis (ORA) to identify potential mechanisms of action related to these proteins, where the biological pathways were ranked by their statistical significance. **D**: The most relevant of the potential pathways were compared to known COVID-19 dysregulated pathways from the literature and the AOP-Wiki database.

### Endocrine-disrupting chemical dataset

A list of 34 commonly used substances known or suspected to act as EDCs was established, based on knowledge from three data sources: the endocrine disruptor assessment list from ECHA (https://echa.europa.eu/fr/ed-assessment, as of April 24, 2020), the one from NIEHS (https://www.niehs.nih.gov/health/topics/agents/endocrine/index.cfm as of April 28, 2020), and the TEDX database (https://endocrinedisruption.org/interactive-tools/tedx-list-of-potential-endocrine-disruptors/search-the-tedx-list, as of April 24,2020).

To explore as much as possible the chemical diversities, EDCs for this study were further selected to represent different chemical classes (Table 1). The CAS numbers were used for data integration.

**Table 1.** List of the 34 major substances known or suspected to be endocrine-disrupting chemicals.

| CAS | chemical.name | abbreviation | CAS | chemical.name | abbreviation |
| --- | --- | --- | --- | --- | --- |
| 35065-27-1 | 2,2',4,4',5,5'-Hexachlorobiphenyl | PCB-153 | 446-72-0 | genistein | - |
| 1746-01-6 | 2,3,7,8- Tetrachlorodibenzodioxin | TCDD | 3194-55-6 | hexabromocyclododecane | HBCD |
| 1912-24-9 | atrazine | - | 118-74-1 | Hexachlorobenzene | HCB |
| 131-56-6 | benzophenone-1 | - | 138261-41-3 | imidacloprid | - |
| 117-81-7 | Bis (2-ethylhexyl)phthalate | DEHP | 625-45-6 | methoxyacetic acid | MAA |
| 620-92-8 | bisohenol F | BPF | 99-76-3 | methyl-paraben | MEPA |
| 80-05-7 | bisphenol A | BPA | 68412-53-3 | nonylphenol ethoxylate | NPEO |
| 80-09-1 | bisphenol S | BPS | 103-90-2 | acetaminophen | - |
| 94-26-8 | butyl-paraben | BUPA | 68631-49-2 | PBDE-153 | PBDE-153 |
| 57-74-9 | Chlordane | - | 5436-43-1 | PBDE-47 | PBDE-47 |
| 2921-88-2 | chlorpyrifos | CPF | 14797-73-0 | perchlorate | - |
|  |  |  |  | Perfluorooctane sulfonic |  |
| 210880-92-5 | Clothianidin | - | 1763-23-1 | acid | PFOS |
| 52315-07-8 | cypermethrin | - | 335-67-1 | Perfluorooctanoic acid | PFOA |
| 486-66-8 | daidzein | - | 67747-09-5 | prochloraz | - |
| 84-74-2 | Dibutyl phthalate | DBP | 153719-23-4 | thiamethoxam | - |
| 72-55-9 | Dichlorodiphenyldichloroethylene | DDE | 688-73-3 | Tributyltin | TBT |
| 50-29-3 | dichlorodiphenyltrichloroethane | DDT | 3380-34-5 | triclosan | - |

**Table 2.**
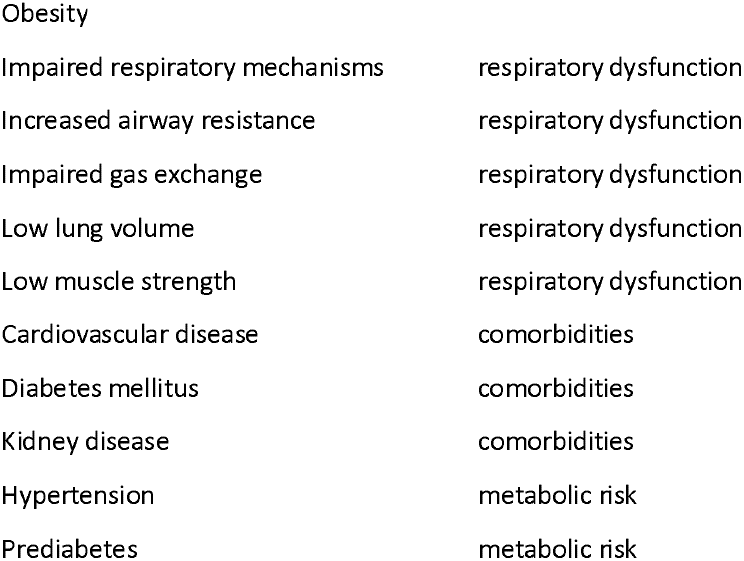
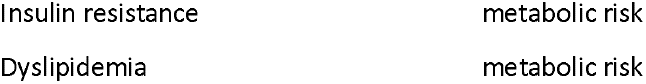
List of the 13 diseases.

### Disease dataset

Comorbidities known to be associated with obesity or otherwise leading to severe COVID-19 were extracted from a recent study (Stefan et al. 2020), and resulted in a total of 13 disorders for exploration in the integrative systems toxicology (Table2).

### Endocrine-disrupting chemical-protein associations

Human proteins known to be associated with each of the 34 EDCs were extracted from the U.S. Environmental Protection Agency web-based CompTox Chemistry dashboard, which contains a wide range of data related to chemical toxicity, including *in vitro* bioassays data (as of April 30, 2020) (Williams et al. 2017). Each linked protein was matched to a gene symbol and classified using the Panther (protein analysis through evolutionary relationships) classification system (version 15, released February 14, 2020) (Mi et al. 2013), a curated biological database of gene/protein families, and their functionally related subfamilies that can be used to classify and identify the function of gene products.

### Disease-protein associations

From two human protein-disease databases, proteins known to be linked to the 13 studied diseases were listed (as of April 29, 2020 for both data sources). The DisGenNet database is a discovery platform containing one of the largest publicly available collections of genes and variants associated with human diseases(Piñero et al. 2015). The GeneCards database contains manually curated information for substances and their associations to genes and proteins, that are scored (Safran et al. 2010). For the present study, only associations were kept only for those between human diseases and proteins categorized as coding proteins, and all non-human information, including gene clusters, genetic locus, pseudogenes, RNA genes and those uncategorized were disregarded. All listed proteins were matched to their gene symbol to facilitate further analysis. Each identified protein from both databases, was categorized into the protein class using the Panther classification (version 15).

### Pathways enrichment analysis

To decipher biological pathways potentially linked to the selected EDCs and explore if they might overlap with the ones known for COVID-19, an ORA was done. Four major sources of protein-pathway information were independently integrated, i.e., using the Kyoto Encyclopedia of Genes and Genomes (KEGG), the Reactome, the Wiki-pathways and the Panther databases(Fabregat et al. 2018; Kanehisa et al. 2019; Mi et al. 2013; Slenter et al. 2018). To assess the statistical significance of the protein-pathway relationships, a hypergeometric test was used for each of the four sources, followed by a multiple testing correction of the *p*-values with the Benjamini-Hochberg method. The ORA was performed on the common proteins identified to identify the most strongly linked proteins that are affected by the EDCs and also associated with at least one the 13 comorbidities. As a last step, manual curation allowed us to consider relevant outcomes for interpretation. The four data sources provided complementary information, with some overlapping findings.

### COVID-19 and biological mechanism of action

Linkage between COVID-19 and potential biological targets and affected pathways were extracted from the literature (as of May 22, 2020) and the AOP-Wiki database (as of May 22, 2020).

## Results

### Endocrine-disrupting chemical-protein associations

From the CompTox database, information on the links between chemicals and human proteins were compiled. Data for 30 of the 34 chemicals could be retrieved, and a total of 208 unique human proteins were involved via 1632 associations. No information was retrieved for hexachlorobenzene, nonylphenol ethoxylate, perchlorate and tributyltin. Perfluorooctane sulfonic acid (PFOS) targeted the highest number of proteins (113), and 2,2’,4,4’,5,5’-hexachlorobiphenyl (PCB 153) was associated with only one biological target (the progesterone receptor). The most frequently affected proteins included the androgen receptor (AR) and the estrogen receptor-alpha (ESR1), which were each linked to 23 EDCs, whereas 61 individual proteins were associated with only one EDC.

To identify the biological targets that are most often affected by EDCs, proteins were grouped in clusters according to their families, as based on the Panther classification system (Figure 2). The majority of the 208 proteins related to EDCs belonged to 12 classes among the 23 present in Panther, while the remaining proteins were classified as ‘uncategorized protein class’. Each protein was assigned to only one category, although only one of them, HLA-DRA, (HLA class II histocompatibility antigen, DR alpha chain) belonged to the defense/immunity group. Other immunity-related proteins. such as interleukin 6 (IL-6) or interleukin 1 alpha (IL-1A), were not associated with any class in the Panther classification. We therefore manually added all immune system-related proteins to the “uncategorized class”. Given that Bisphenol A (BPA) increases the release of these proteins(Ben-Jonathan et al. 2009), and because antibodies to the IL-6 receptor (such as tocilizumab) or to the IL-1 receptor (such as Anakinra) are currently tested for the treatment of COVID-19 patients (Zhou et al. 2020), we also explored if the proteins selected could be mapped to defense and/or immunity biological categories. For this purpose, we used the Gene Ontology (GO) classification (as of May 26, 2020), and among the 208 proteins dysregulated by EDCs, 58 were associated with inflammatory response, 75 with defense response, and 66 with regulation of immune system process.

**Figure 2.**
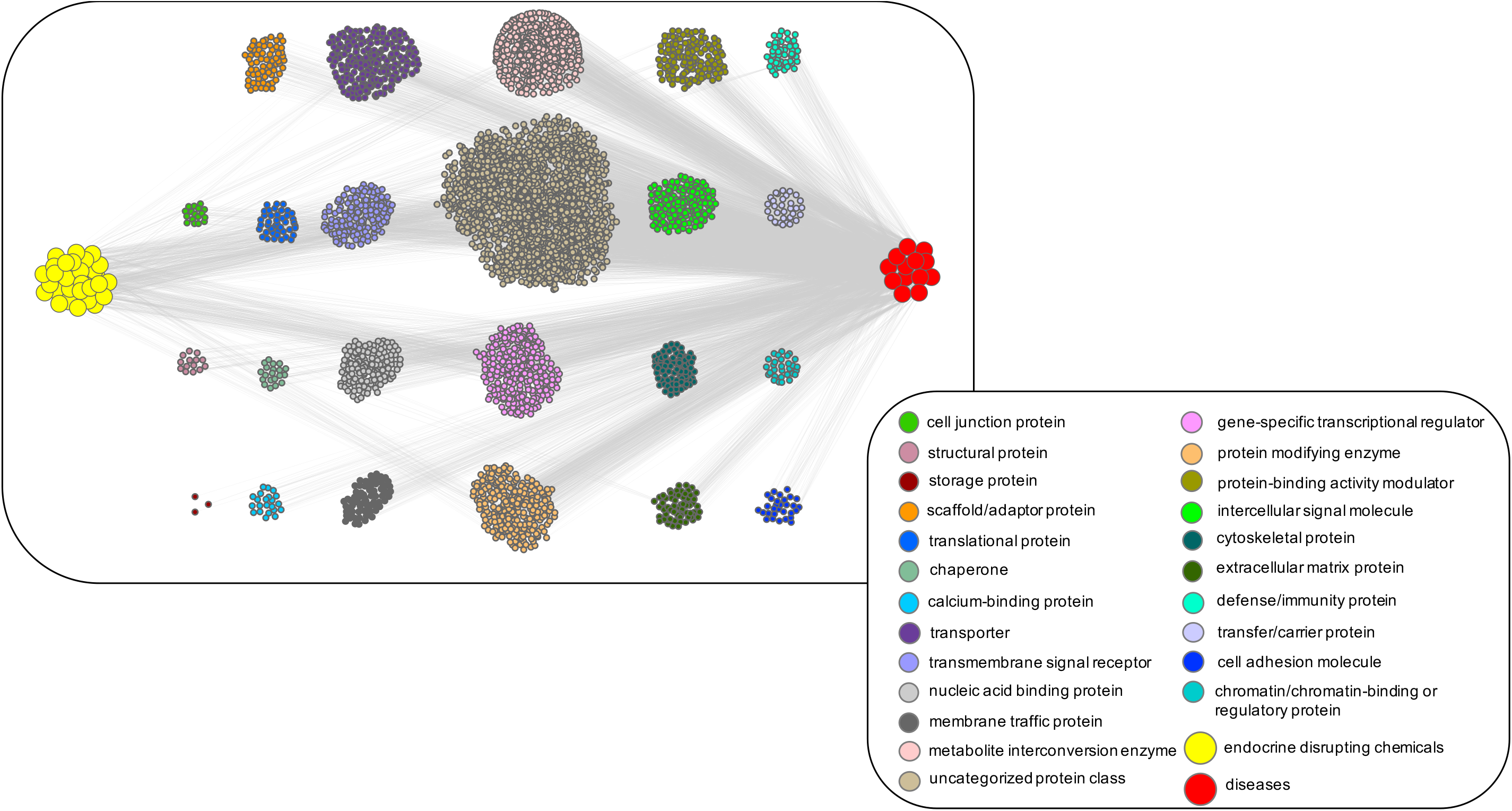
Tripartite network representation of endocrine-disrupting chemicals-proteins-diseases relationships. First, a bipartite network of the 208 human proteins known to be dysregulated by the 30 endocrine-disrupting chemicals (EDCs) was created as extracted from the CompTox database. Each yellow diamond node represents an EDC, and edges are the interactions between EDCs and proteins. Then, a second bipartite network was generated for the 4184 human proteins known to be linked to the 13 predisposing diseases, as extracted from the DisGeNET (3262 links) and GeneCards (2079 links) databases. Each red square node represents a disease, and edges are the interactions between diseases and proteins. A total of 1156 proteins were overlapping. All proteins were grouped using the Panther classification system (version 15) and are represented by circles (colors are according to their Panther family classes).

### Disease-protein associations

Regarding diseases associated with human proteins, two databases were screened. From the DisGeNET database, we were able to retrieve information for 8 of the 13 diseases, which were connected to 3262 unique proteins via 7195 links (as of April 29, 2020). The proteins were categorized in 22 protein classes using the Panther classification (version 15) (Figure S1). Proteins that did not belong in any class were again grouped into the uncategorized class. Obesity and diabetes were linked to proteins belonging to each of the 22 categories, whereas insulin resistance and dyslipidemia were linked to only half of the categories.

From the GeneCards database, all 13 predisposing diseases were retrieved (as of 29 April 2020), and a total of 115,289 associations were identified between the diseases and 29,094 unique human proteins were extracted. Among them, only protein-coding information according to HGNC, Ensembl or Entrez Gene were kept (proteins data related to biological regions, gene clusters, genetic loci, pseudogenes, non-coding RNA genes and uncategorized elements were not considered), thereby reducing the total number of unique protein to 18,931, representing 97,855 disease-protein links. As a next step, grouping of the proteins using the Panther classification system allowed identification of 23 clusters correspond to the 23 different protein classes (Figure S2). Each protein was assigned to only one category, except for ameloblastin (AMBN), which was associated with both ‘extracellular matrix protein’ and ‘structural protein’. Proteins not associated with Panther classes, were again grouped into the uncategorized class. Excluding the viral or transposable element protein class, all diseases (except dyslipidemia) were associated with all the other Panther classes.

In order to keep the most relevant protein-disease associations obtained from the GeneCards database, data were filtered based on their scores. The GeneCards scores are calculated based on publications mentioning a protein and a disease, using a Boolean model. The higher the score, the more relevant the protein-disease association is. Among the 97,855 links between the 13 diseases and 18,931 proteins, the score values ranged between 0.13 (representing very low association) to 228 (very high evidence for a protein-disease connection). After evaluation of the extracted data (number of proteins by GeneCards scores), we selected associations with a score ≥ 20 (see Figure S3). Within this threshold, a total of 5732 associations were retained that link the 12 diseases with 2079 unique human proteins (no information was retained for ‘dyslipidemias’ from the GeneCards database).

### Generating a tripartite network of protein-EDC-disease associations

A human bipartite associative network of proteins and the 13 diseases was created. Among the 3262 unique proteins from the DisGeNET, and the 2079 proteins from the GeneCards databases, 1157 were overlapping proteins and only 922 and 2105 proteins were uniquely associated with GeneCards or DisGeNET, respectively. All 4184 unique proteins were again grouped into 23 clusters using the Panther classification (the class ‘viral or transposable element protein’ was not kept after the cleaning step. Among the groupings, we retrieved a cluster of proteins linked to the ‘defense/immunity’ category. These results were merged with the bipartite protein-EDCs network to develop a tripartite network (Figure 2).

### Translation into pathways

To identify biological pathways that may be involved in the predisposing diseases while also being dysregulated by the EDCs, we first analyzed the overlaps between the two sets of proteins. Among the proteins identified from the three data sources, 98 were common (Figure S4), and all of them were mapped to unique Entrez GeneID, and could therefore be used for biological enrichment analyses, which were performed independently using four data sources (KEGG, Reactome, Wiki-pathways and Panther). The ORA analysis revealed several statistically significant pathways linked to interleukins/cytokines signaling, intracellular signaling pathways and, regulation of metabolic pathways (Table 3). Interestingly, the different data sources showed very significant associations with common pathways, such as interleukins (IL) related pathways: IL-4 and IL-13, IL-10 signaling pathways (p_adj_ < E-16, and p_adj_ of 2.85E-09 respectively, Reactome), IL-17 signaling pathway (p_adj_ of 1.05E-10, KEGG), IL-3, IL-5 and IL-18 signaling pathways (p_adj_ of 1.09E-09, 2.52E-09, 1.49E-08 respectively, Wiki-pathways), the IL-signaling pathways (p_adj_ of 1.10E-05, Panther); or the Toll-like receptor signaling pathway (p_adj_ 3.91E-09 for KEGG, p_adj_ of 0.99 for Panther and p_adj_ 1.93E-08 for Wiki-pathways).

**Table 3.**
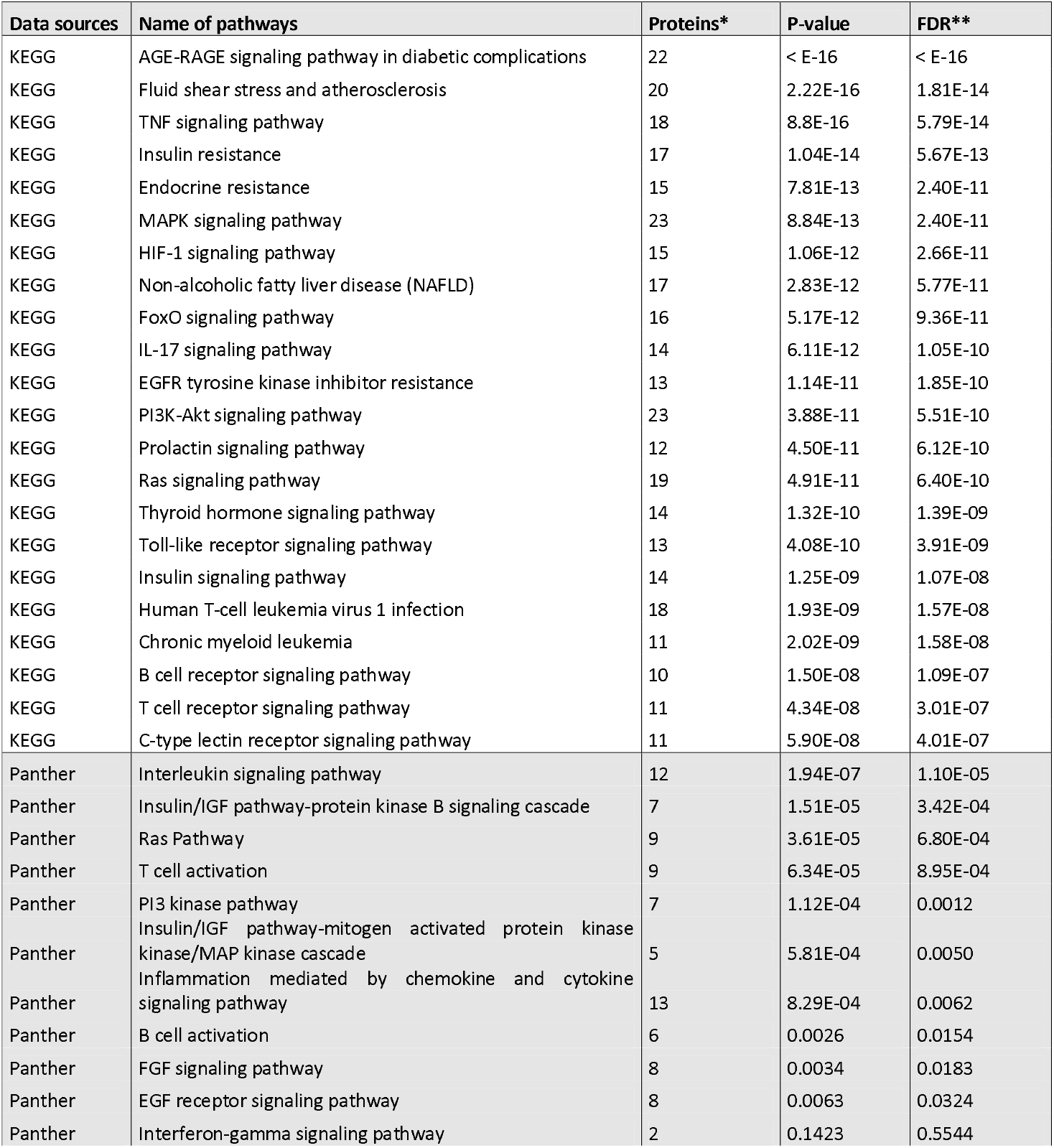
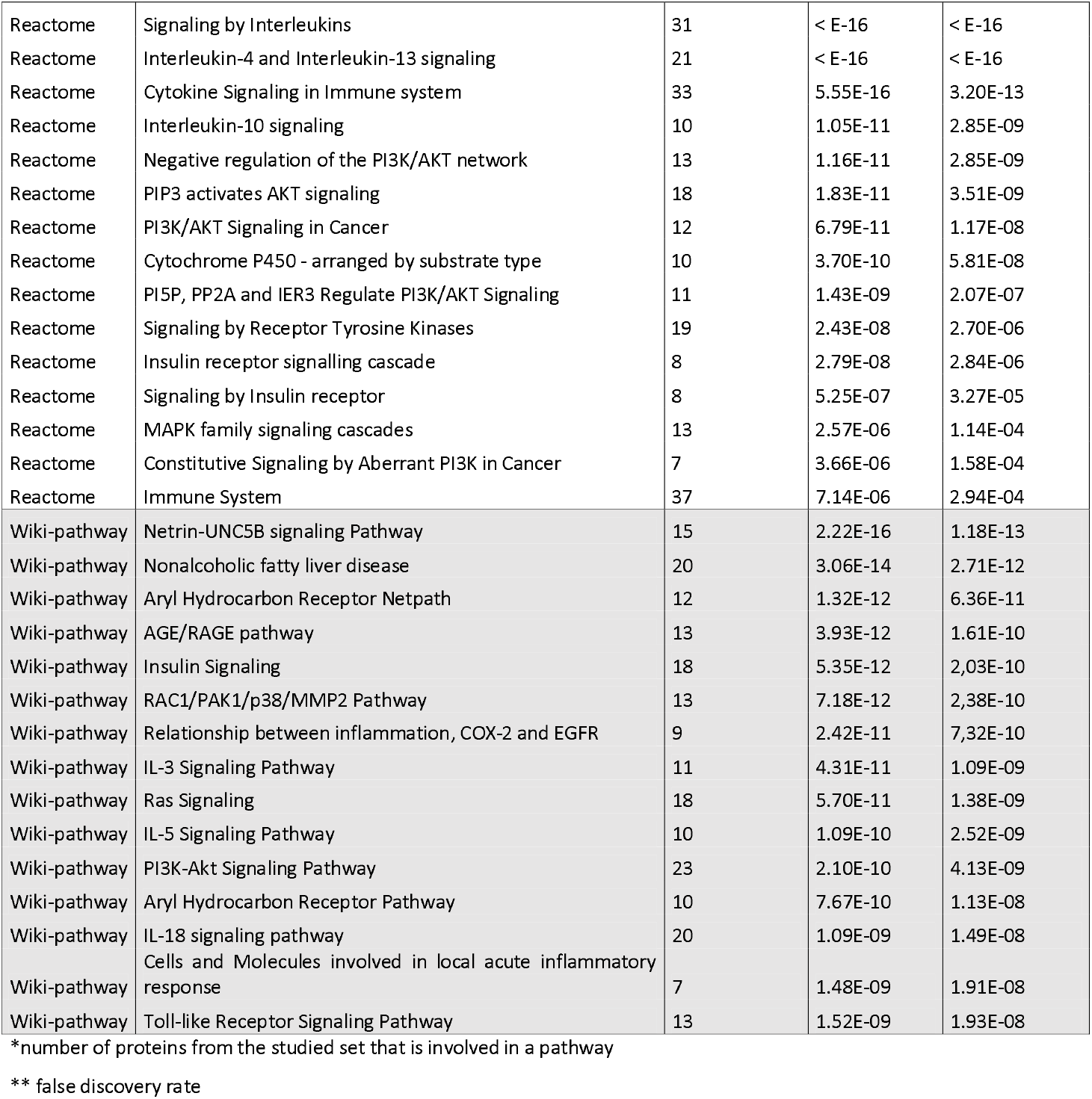
Pathway enrichment for the set of proteins that are linked to both the predisposing diseases and to the EDCs. The pathways were extracted from the KEGG, Panther, Reactome and the Wikipathways database.

Among the most significant pathways, several were retrieved from each of the data sources with relation to the AGE/RAGE pathway (*i.e*. Advanced Glycation End products and its receptor), which is known to cause cellular stress and inflammation. The AGE are formed non-enzymatically, by Maillard reaction products (carbohydrates with proteins and/or lipids) and bind to the RAGE. Formation of AGE has been associated with chronic diseases such as type 2 diabetes (Cai et al. 2012; Menini et al. 2018). Similarly, the stress or inflammatory pathways (e.g shear stress, defined as the tangential force exerted by the blood flow on the vascular endothelium, TNF-alpha) are highlighted by our analysis; the shear stress activates the AhR signaling pathway, which is also involved in the regulation of IL-17 production by the Th17 lymphocytes; interleukin 17 has been suspected to be involved in the pathogenesis of COVID19(Gutiérrez-Vázquez and Quintana 2018; Han et al. 2008; Pacha et al. 2020). Interestingly, inflammation is suspected to influence insulin resistance.

### Exploration of EDCs linkage to COVID-19

To explore putative links between COVID-19 and exposure to EDCs, we first screened the AOP-Wiki database, and then further examined the pathways identified using literature references.

In the AOP-Wiki database, only one AOP was related to COVID-19, and it involves several key events, such as ‘increased pro-inflammatory mediators’ (KE 1496), ‘increased inflammatory immune responses’ (KE 1750), which leads to the adverse outcome ‘increased mortality’ (AO 351). Such knowledge-based linear chain of events highlights the importance of the link between COVID-19 and inflammatory processes.

## Discussion

In order to investigate possible links between exposure to EDCs and the severity of COVID-19, we explored a computational systems biology approach. The tripartite network model first linked EDCs to targeted proteins and then proteins related to diseases that predispose to more serious COVID-19 development, thereby allowing us to identify common signaling pathways. The identification of such joint pathways and their role as possible targets of EDCs highlights the potential links between exposure to environmental chemicals and COVID-19 severity.

This integrative approach can be easily applied as a new approach methodology (NAM) (Bopp et al. 2019), which may offer support to methods alternative to animal testing or to identify biological pathways that require more focused laboratory study. Previous studies have demonstrated that systems chemical toxicology models combined with computational network biology may help in understanding chemical toxicity in humans (Hartung et al. 2017; Nie et al. 2015; Taboureau and Audouze 2017). Our tripartite network supports the notion that exposure to EDCs may contribute to aggravation of COVID-19. Although major links were identified at extremely low p values, the approach relies on existing information available in within the very substantive data sources, but some causal associations may have been overlooked or disregarded because of missing or incomplete information.

To assess the validity of our approach, a more focused expert analysis was attempted, where we selected the Th17 and the AGE/RAGE signaling pathways because of their pathophysiological relevance in the context of COVID-19. The interleukin-17 (IL-17) signaling pathway plays several important roles, and IL-17 is produced by a pro-inflammatory subtype of T helper lymphocytes named Th17 cells, located at mucosal barriers where they contribute to pathogen clearance. The IL-17 produced stimulates the synthesis of cytokines (IL1ß, TNF-alpha…) and chemokines (MCP-1…) by other cell types, thereby favoring the recruitment of monocytes and neutrophils at inflammatory sites. However, an over-activation of Th17 cells can lead to a hyper-inflammatory state which is deleterious (Pacha et al. 2020).

The highly variable symptomatology associated with the infection by SARS-CoV-2 depends on the levels of IL-17 and of other cytokines including IL-1ß, IL-6, IL-15, TNF-alpha and IFNγ. The most deleterious effect of SARS-CoV-2 in humans is an acute lung injury leading to a severe acute respiratory syndrome (SARS) that is partly due to IL-17-related excessive recruitment of pro-inflammatory cells and production of pro-inflammatory cytokines. Therefore, an increased basal level of IL-17 (in the absence of infection, for example due to obesity or to induction by a chemical) might represent a lung injury risk associated with SARS-CoV-2 infection. Our finding of EDC linkage to this pathway is therefore of high pathogenetic relevance.

Obesity promotes a high basal level of inflammation which contributes to insulin resistance and type 2 diabetes(Goldberg 2009). This phenomenon is due to an infiltration of the adipose tissue (AT) by macrophages and T cells and their production of various pro-inflammatory cytokines, including IL-1ß, TNF-alpha, IL-17 and IL-6. Several EDCs are suspected to be obesogenic (and are subsequently named obesogens). This has been demonstrated for several substances (e.g. tributyltin) and linked to the stimulation of pro-adipogenic signaling pathway (e.g. through PPARγ)(Egusquiza and Blumberg 2020). Similarly, the aryl hydrocarbon receptor (AhR) is highly expressed in Th17 cells and is an essential contributor to the production of IL-17(Veldhoen et al. 2008). The AhR, known as the receptor of dioxins and dioxin-like PCBs, is also activated by shear stress (SS), another pathway highlighted in our computational analysis. Indeed, several studies have shown using various endothelial models that laminar SS leads to the activation of two target genes of the AhR, namely CYP1A1 and CYP1B1(Conway et al. 2009). Two recent studies suggest an indirect link between SARS-CoV-2 and SS by showing that the expression of ACE2 (angiotensin-converting enzyme 2), the receptor of the virus, is increased by SS (Song et al. 2020).

These observations support a dual impact of EDCs on IL-17 production and inflammatory state; this impact could be indirect due to the effect of these chemicals on obesity or through a direct stimulation of several signaling pathways, such as AhR or PPARγ, leading to an overproduction of systemic IL-17; the shear stress pathway represents an additional link between AhR activation and the EDC/disease connection. The implication of shear stress also suggests a possible contribution of increased expression of ACE2, the receptor of the SARS-CoV-2. While the role of these pathways at the nexus between exposure to EDCs and COVID-19 severity appears to be relevant, their actual contribution remains to be demonstrated and their putative role as therapeutic targets remains to be further substantiated.

Our integrative systems biology study also indicates a strong statistical association between the AGE/RAGE signaling pathway, chronic diseases and EDC effects. This is likely due to the well-known links between this pathway and type 2 diabetes(Ravichandran et al. 2019). Indeed, hyperglycemia leads to increased amounts of glycation products and their metabolites which results in the activation of the RAGE receptors. The latter are highly expressed in endothelial cells, and their activation leads to increased oxidative stress and inflammation and ultimately to endothelial damage, thrombotic disorders and vascular diseases (Egaña-Gorroño et al. 2020). Other endogenous ligands can also activate RAGE, among them HMGB1 (high-mobility group box 1), an extra-cellular protein also linked to a variety of inflammatory responses(Andersson et al. 2020). Interestingly, the AGE/RAGE signaling pathway is highly expressed in the lung vasculature and has been implicated in several pulmonary diseases(Oczypok et al. 2017). All these observations support the implication of the AGE/RAGE signaling pathway in vascular, thrombotic and lung diseases which are the hallmarks of COVID-19 severity. Interestingly, there are also complex connections between HMGB1 and ACE2 which is the receptor for SARS-Cov2 and other coronaviruses(Luft 2016). These results are in accordance with recent proposals in published commentaries of environmental chemical impacts on COVID-19 progress(Andersson et al. 2020; Rojas et al. 2020).

The three-way approach did not attempt to identify direct immunotoxic effects due to environmental chemicals otherwise considered to be EDCs. However, some of the EDCs selected, i.e., PCB-153, PFOA and PFOS, are known to have immunotoxic properties (Heilmann et al.), and the same is true for some common air pollutants (Tsatsakis et al. 2020). Accordingly, the impact of environmental chemicals on COVID-19 severity demands attention.

## Conclusions

The results of this computational study appear as a promising initial step toward systematically linking a major group of environmental chemicals to the severity of COVID-19, although the findings need to be further supported by high-throughput screening tests, clinical and experimental data. Nevertheless, these observations bridge environmental stressors and infectious diseases and support an integrated exposome approach. Preliminary focus on the AGE/RAGE and IL-17 pathways illustrates the potential connection between exposure to EDCs and diseases predisposing to COVID-19 severity.

## Data Availability

The authors confirm that the data supporting the findings of this study are available within the article [and/or] its supplementary materials.

## Acknowledgments

The authors would like to acknowledge OBERON (https://oberon-4eu.com/), a project funded by the European Union’s Horizon 2020 research and innovation program under grant agreement no. 825712. This work was also supported by the University of Paris and INSERM. PG is supported by the National Institute of Environmental Health Sciences, NIH (P42ES027706)

## References

1. Andersson U, Ottestad W, Tracey KJ. 2020. Extracellular HMGB1: a therapeutic target in severe pulmonary inflammation including COVID-19? Mol Med Camb Mass 26:42; doi:10.1186/s10020-020-00172-4.

2. Audouze K, Brunak S, Grandjean P. 2013. A computational approach to chemical etiologies of diabetes. Sci Rep 3:2712; doi:10.1038/srep02712.

3. Audouze K, Taboureau O, Grandjean P. 2018. A systems biology approach to predictive developmental neurotoxicity of a larvicide used in the prevention of Zika virus transmission. Toxicol Appl Pharmacol 354:56–63; doi:10.1016/j.taap.2018.02.014.

4. Bashir MF, Ma BJ, Bilal null, Komal B, Bashir MA, Farooq TH, et al. 2020. Correlation between environmental pollution indicators and COVID-19 pandemic: A brief study in Californian context. Environ Res 187:109652; doi:10.1016/j.envres.2020.109652.

5. Ben-Jonathan N, Hugo ER, Brandebourg TD. 2009. Effects of bisphenol A on adipokine release from human adipose tissue: Implications for the metabolic syndrome. Mol Cell Endocrinol 304:49–54; doi:10.1016/j.mce.2009.02.022.

6. Bopp SK, Kienzler A, Richarz A-N, van der Linden SC, Paini A, Parissis N, et al. 2019. Regulatory assessment and risk management of chemical mixtures: challenges and ways forward. Crit Rev Toxicol 49:174–189; doi:10.1080/10408444.2019.1579169.

7. Bornstein SR, Dalan R, Hopkins D, Mingrone G, Boehm BO. 2020. Endocrine and metabolic link to coronavirus infection. Nat Rev Endocrinol 16:297–298; doi:10.1038/s41574-020-0353-9.

8. Cai W, Ramdas M, Zhu L, Chen X, Striker GE, Vlassara H. 2012. Oral advanced glycation endproducts (AGEs) promote insulin resistance and diabetes by depleting the antioxidant defenses AGE receptor-1 and sirtuin 1. Proc Natl Acad Sci U S A 109:15888–15893; doi:10.1073/pnas.1205847109.

9. Cipelli R, Harries L, Okuda K, Yoshihara S, Melzer D, Galloway T. 2014. Bisphenol A modulates the metabolic regulator oestrogen-related receptor-α in T-cells. Reprod Camb Engl 147:419– 426; doi:10.1530/REP-13-0423.

10. Conway DE, Sakurai Y, Weiss D, Vega JD, Taylor WR, Jo H, et al. 2009. Expression of CYP1A1 and CYP1B1 in human endothelial cells: regulation by fluid shear stress. Cardiovasc Res 81:669–677; doi:10.1093/cvr/cvn360.

11. Couleau N, Falla J, Beillerot A, Battaglia E, D’Innocenzo M, Plançon S, et al. 2015. Effects of Endocrine Disruptor Compounds, Alone or in Combination, on Human Macrophage-Like THP-1 Cell Response. PloS One 10:e0131428; doi:10.1371/journal.pone.0131428.

12. Dalsager L, Christensen N, Husby S, Kyhl H, Nielsen F, Høst A, et al. 2016. Association between prenatal exposure to perfluorinated compounds and symptoms of infections at age 1-4years among 359 children in the Odense Child Cohort. Environ Int 96:58–64; doi:10.1016/j.envint.2016.08.026.

13. Drucker DJ. 2020. Coronavirus Infections and Type 2 Diabetes-Shared Pathways with Therapeutic Implications. Endocr Rev 41; doi:10.1210/endrev/bnaa011.

14. Egaña-Gorroño L, López-Díez R, Yepuri G, Ramirez LS, Reverdatto S, Gugger PF, et al. 2020. Receptor for Advanced Glycation End Products (RAGE) and Mechanisms and Therapeutic Opportunities in Diabetes and Cardiovascular Disease: Insights From Human Subjects and Animal Models. Front Cardiovasc Med 7:37; doi:10.3389/fcvm.2020.00037.

15. Egusquiza RJ, Blumberg B. 2020. Environmental Obesogens and Their Impact on Susceptibility to Obesity: New Mechanisms and Chemicals. Endocrinology 161; doi:10.1210/endocr/bqaa024.

16. Fabregat A, Jupe S, Matthews L, Sidiropoulos K, Gillespie M, Garapati P, et al. 2018. The Reactome Pathway Knowledgebase. Nucleic Acids Res 46:D649–D655; doi:10.1093/nar/gkx1132.

17. Fattorini D, Regoli F. 2020. Role of the chronic air pollution levels in the Covid-19 outbreak risk in Italy. Environ Pollut Barking Essex 1987 264:114732; doi:10.1016/j.envpol.2020.114732.

18. Goldberg RB. 2009. Cytokine and cytokine-like inflammation markers, endothelial dysfunction, and imbalanced coagulation in development of diabetes and its complications. J Clin Endocrinol Metab 94:3171–3182; doi:10.1210/jc.2008-2534.

19. Grandjean P, Andersen EW, Budtz-Jørgensen E, Nielsen F, Mølbak K, Weihe P, et al. 2012. Serum vaccine antibody concentrations in children exposed to perfluorinated compounds. JAMA 307:391–397; doi:10.1001/jama.2011.2034.

20. Granum B, Haug LS, Namork E, Stølevik SB, Thomsen C, Aaberge IS, et al. 2013. Pre-natal exposure to perfluoroalkyl substances may be associated with altered vaccine antibody levels and immune-related health outcomes in early childhood. J Immunotoxicol 10:373– 379; doi:10.3109/1547691X.2012.755580.

21. Gutiérrez-Vázquez C, Quintana FJ. 2018. Regulation of the Immune Response by the Aryl Hydrocarbon Receptor. Immunity 48:19–33; doi:10.1016/j.immuni.2017.12.012.

22. Han Z, Miwa Y, Obikane H, Mitsumata M, Takahashi-Yanaga F, Morimoto S, et al. 2008. Aryl hydrocarbon receptor mediates laminar fluid shear stress-induced CYP1A1 activation and cell cycle arrest in vascular endothelial cells. Cardiovasc Res 77:809–818; doi:10.1093/cvr/cvm095.

23. Hartung T, FitzGerald RE, Jennings P, Mirams GR, Peitsch MC, Rostami-Hodjegan A, et al. 2017. Systems Toxicology: Real World Applications and Opportunities. Chem Res Toxicol 30:870–882; doi:10.1021/acs.chemrestox.7b00003.

24. Heilmann C, Grandjean P. Immunotoxicity: Impacts and Research Approaches. In: Kishi R, Grandjean P, editors. Health Impacts of Developmental Exposure to Environmental Chemicals. Singapore: Springer; 2020. pp. 175–90.

25. Kanehisa M, Sato Y, Furumichi M, Morishima K, Tanabe M. 2019. New approach for understanding genome variations in KEGG. Nucleic Acids Res 47:D590–D595; doi:10.1093/nar/gky962.

26. Luft FC. 2016. High-mobility group box 1 protein, angiotensins, ACE2, and target organ damage. J Mol Med Berl Ger 94:1–3; doi:10.1007/s00109-015-1372-1.

27. Menini S, Iacobini C, de Latouliere L, Manni I, Ionta V, Blasetti Fantauzzi C, et al. 2018. The advanced glycation end-product N1 -carboxymethyllysine promotes progression of pancreatic cancer: implications for diabetes-associated risk and its prevention. J Pathol 245:197–208; doi:10.1002/path.5072.

28. Mi H, Muruganujan A, Thomas PD. 2013. PANTHER in 2013: modeling the evolution of gene function, and other gene attributes, in the context of phylogenetic trees. Nucleic Acids Res 41:D377–386; doi:10.1093/nar/gks1118.

29. Nie W, Lv Y, Yan L, Chen X, Lv H. 2015. Prediction and Characterisation of the System Effects of Aristolochic Acid: A Novel Joint Network Analysis towards Therapeutic and Toxicological Mechanisms. Sci Rep 5:17646; doi:10.1038/srep17646.

30. Oczypok EA, Perkins TN, Oury TD. 2017. All the “RAGE” in lung disease: The receptor for advanced glycation endproducts (RAGE) is a major mediator of pulmonary inflammatory responses. Paediatr Respir Rev 23:40–49; doi:10.1016/j.prrv.2017.03.012.

31. Pacha O, Sallman MA, Evans SE. 2020. COVID-19: a case for inhibiting IL-17? Nat Rev Immunol 20:345–346; doi:10.1038/s41577-020-0328-z.

32. Petrilli CM, Jones SA, Yang J, Rajagopalan H, O’Donnell L, Chernyak Y, et al. 2020. Factors associated with hospital admission and critical illness among 5279 people with coronavirus disease 2019 in New York City: prospective cohort study. BMJ 369:m1966; doi:10.1136/bmj.m1966.

33. Piñero J, Queralt-Rosinach N, Bravo À, Deu-Pons J, Bauer-Mehren A, Baron M, et al. 2015. DisGeNET: a discovery platform for the dynamical exploration of human diseases and their genes. Database J Biol Databases Curation 2015:bav028; doi:10.1093/database/bav028.

34. Ravichandran G, Lakshmanan DK, Raju K, Elangovan A, Nambirajan G, Devanesan AA, et al. 2019. Food advanced glycation end products as potential endocrine disruptors: An emerging threat to contemporary and future generation. Environ Int 123:486–500; doi:10.1016/j.envint.2018.12.032.

35. Rojas A, Gonzalez I, Morales MA. 2020. SARS-CoV-2-mediated inflammatory response in lungs: should we look at RAGE? Inflamm Res Off J Eur Histamine Res Soc Al 69:641–643; doi:10.1007/s00011-020-01353-x.

36. Safran M, Dalah I, Alexander J, Rosen N, Iny Stein T, Shmoish M, et al. 2010. GeneCards Version 3: the human gene integrator. Database J Biol Databases Curation 2010:baq020; doi:10.1093/database/baq020.

37. Slenter DN, Kutmon M, Hanspers K, Riutta A, Windsor J, Nunes N, et al. 2018. WikiPathways: a multifaceted pathway database bridging metabolomics to other omics research. Nucleic Acids Res 46:pD661–D667; doi:10.1093/nar/gkx1064.

38. Song J, Hu B, Qu H, Wang L, Huang X, Li M, et al. 2020. Upregulation of angiotensin converting enzyme 2 by shear stress reduced inflammation and proliferation in vascular endothelial cells. Biochem Biophys Res Commun 525:812–818; doi:10.1016/j.bbrc.2020.02.151.

39. Stefan N, Birkenfeld AL, Schulze MB, Ludwig DS. 2020. Obesity and impaired metabolic health in patients with COVID-19. Nat Rev Endocrinol; doi:10.1038/s41574-020-0364-6.

40. Taboureau O, Audouze K. 2017. Human Environmental Disease Network: A computational model to assess toxicology of contaminants. ALTEX 34:289–300; doi:10.14573/altex.1607201.

41. Trasande L, Zoeller RT, Hass U, Kortenkamp A, Grandjean P, Myers JP, et al. 2016. Burden of disease and costs of exposure to endocrine disrupting chemicals in the European Union: an updated analysis. Andrology 4:565–572; doi:10.1111/andr.12178.

42. Tsatsakis A, Petrakis D, Nikolouzakis TK, Docea AO, Calina D, Vinceti M, et al. 2020. COVID-19, an opportunity to reevaluate the correlation between long-term effects of anthropogenic pollutants on viral epidemic/pandemic events and prevalence. Food Chem Toxicol Int J Publ Br Ind Biol Res Assoc 141:111418; doi:10.1016/j.fct.2020.111418.

43. Vandenberg LN, Ågerstrand M, Beronius A, Beausoleil C, Bergman Å, Bero LA, et al. 2016. A proposed framework for the systematic review and integrated assessment (SYRINA) of endocrine disrupting chemicals. Environ Health Glob Access Sci Source 15:74; doi:10.1186/s12940-016-0156-6.

44. Veldhoen M, Hirota K, Westendorf AM, Buer J, Dumoutier L, Renauld J-C, et al. 2008. The aryl hydrocarbon receptor links TH17-cell-mediated autoimmunity to environmental toxins. Nature 453:106–109; doi:10.1038/nature06881.

45. Vermeulen R, Schymanski EL, Barabási A-L, Miller GW. 2020. The exposome and health: Where chemistry meets biology. Science 367:392–396; doi:10.1126/science.aay3164.

46. Williams AJ, Grulke CM, Edwards J, McEachran AD, Mansouri K, Baker NC, et al. 2017. The CompTox Chemistry Dashboard: a community data resource for environmental chemistry. J Cheminformatics 9:61; doi:10.1186/s13321-017-0247-6.

47. Wu Q, Achebouche R, Audouze K. 2020. Computational systems biology as an animal-free approach to characterize toxicological effects of persistent organic pollutants. ALTEX 37:287–299; doi:10.14573/altex.1910161.

48. Zhou F, Yu T, Du R, Fan G, Liu Y, Liu Z, et al. 2020. Clinical course and risk factors for mortality of adult inpatients with COVID-19 in Wuhan, China: a retrospective cohort study. Lancet Lond Engl 395:1054–1062; doi:10.1016/S0140-6736(20)30566-3.

49. Zhu Y, Xie J, Huang F, Cao L. 2020. Association between short-term exposure to air pollution and COVID-19 infection: Evidence from China. Sci Total Environ 727:138704; doi:10.1016/j.scitotenv.2020.138704.

